# Radiation Therapy Quality Assurance (RT QA) of adjuvant fractionated radiotherapy for atypical meningioma in the ROAM/EORTC1308 trial

**DOI:** 10.64898/2026.08.10.26360149

**Authors:** Helen MO Mayles, Brian J Haylock, Gillian Whitfield, Shaveta Mehta, Robert Brass, Alice Brain, Michael D Jenkinson, Damien C Weber, the ROAM/EORTC-1308 trial management group, and the TROG and UK RTTQA groups

**Affiliations:** Clatterbridge Cancer Centre, Liverpool, UK; Christie NHS Foundation Trust, Manchester, UK; University of Liverpool; Centre for Proton Therapy, Paul Scherrer Institute, Villigen, Switzerland

## Abstract

**BACKGROUND AND PURPOSE:** ROAM/EORTC-1308 is an international (Europe and Australasia) phase III randomised controlled trial (RCT) comparing radiotherapy (60Gy/30#) to observation after surgery for atypical meningioma. The treatment plans of all the patients randomised to radiotherapy were reviewed and approved prior to treatment delivery, either by the UK RTTQA group (European sites) or the Australian TROG (Australia and New Zealand). Pretrial credentialling included outlining and planning a benchmark case (DR).

**MATERIALS AND METHODS:** Variations from the guidelines were noted in quality assurance (QA) reports both for DR and for the on-trial Individual Case Reviews (ICRs) and classed as major or minor. After recruitment had finished, a random 10% of the ICRs were independently reviewed for audit purposes. We used the QA reports to analyse the variations in the DR and in the ICRs

**RESULTS:** 56 sites (20 UK, 25 EORTC, 11 TROG) undertook the DR of which 13 (23%) had major variations. During the trial 64 patients at 28 sites received radiotherapy, the median being 2 patients per site. Overall, 25 (39%) patient cases needed resubmitting: 22 (36%) sets of outlines (2 cases twice) and 12 (14%) treatment plans (1 plan twice).

**CONCLUSIONS:** For complex radiotherapy of rare tumours, a DR is insufficient and prospective ICRs of all patients is required. The consistent high-quality radiotherapy in ROAM/EORTC1308 ensures the primary outcome (progression free survival) will be a robust assessment and any difference between treatment arms cannot be attributed to variation in radiotherapy treatment.

## Background and Purpose

Atypical meningioma (WHO grade 2) constitute up to 35% of all meningioma [1] and tend to have a less favourable prognosis than grade 1 meningioma, with an estimated 5-year recurrence rate of 39-58%[2]. The role of adjuvant radiotherapy for reducing the risk of recurrence after gross total resection (Simpson 1-3) for atypical meningioma continues to be disputed and there is no consensus on the optimal treatment in this tumour population [3-8]. Guidelines from the European Association for Neuro-Oncology advise that observation or radiotherapy are both options but do not make a specific recommendation for individual patient management [9]. The ROAM/EORTC-1308 study [3] is the first randomised controlled trial (RCT) of atypical meningioma that compares surgery and adjuvant radiotherapy to surgery and observation after gross total resection (defined as Simpson 1-3 resection [10]).

The primary objective was to determine whether early adjuvant fractionated external beam radiotherapy reduces the risk of tumour recurrence or death due to any cause, compared to active monitoring in newly diagnosed atypical meningioma. The radiotherapy prescription was a median (or mean) dose of 60Gy in 30 daily fractions to the PTV.

In the ROAM trial we wanted to ensure that patients randomised to radiotherapy received treatment according to protocol. We established a prospective Radiotherapy Quality Assurance (RT QA) program coordinated by UK RTTQA, which comprised three consultant clinical / radiation oncologists and three physicists. This group performed the credentialing for all UK and EORTC centres. After reviewing their benchmark cases, it accredited TROG to credential Australian and New Zealand centres. During the trial TROG took full responsibility for the Individual Case Reviews (ICRs). On completion of the trial they returned all the patient DICOM files and QA reviews to the UK RTTQA group.

Pre-trial development meetings, discussions at national and international Neuro-Oncology meetings and an initial literature review, were used to develop the guidelines for radiotherapy contouring and planning (RT guidelines) (Supplementary Appendix B). For contouring they described how to use the Pre-Op MRI to help define the initial extent of the gross tumour volume (GTV), (resection cavity, clearly thickened dural tails and hyperostotic bone) on the Post-Op MRI (Figure 1) and how to use 5mm and 10mm construction lines around the GTV to produce the clinical target volume (CTV), (defined as GTV with a 5mm margin, extending to 10mm along the dura and into brain if there was evidence of brain invasion) (Figure 1).

**Fig 1.**
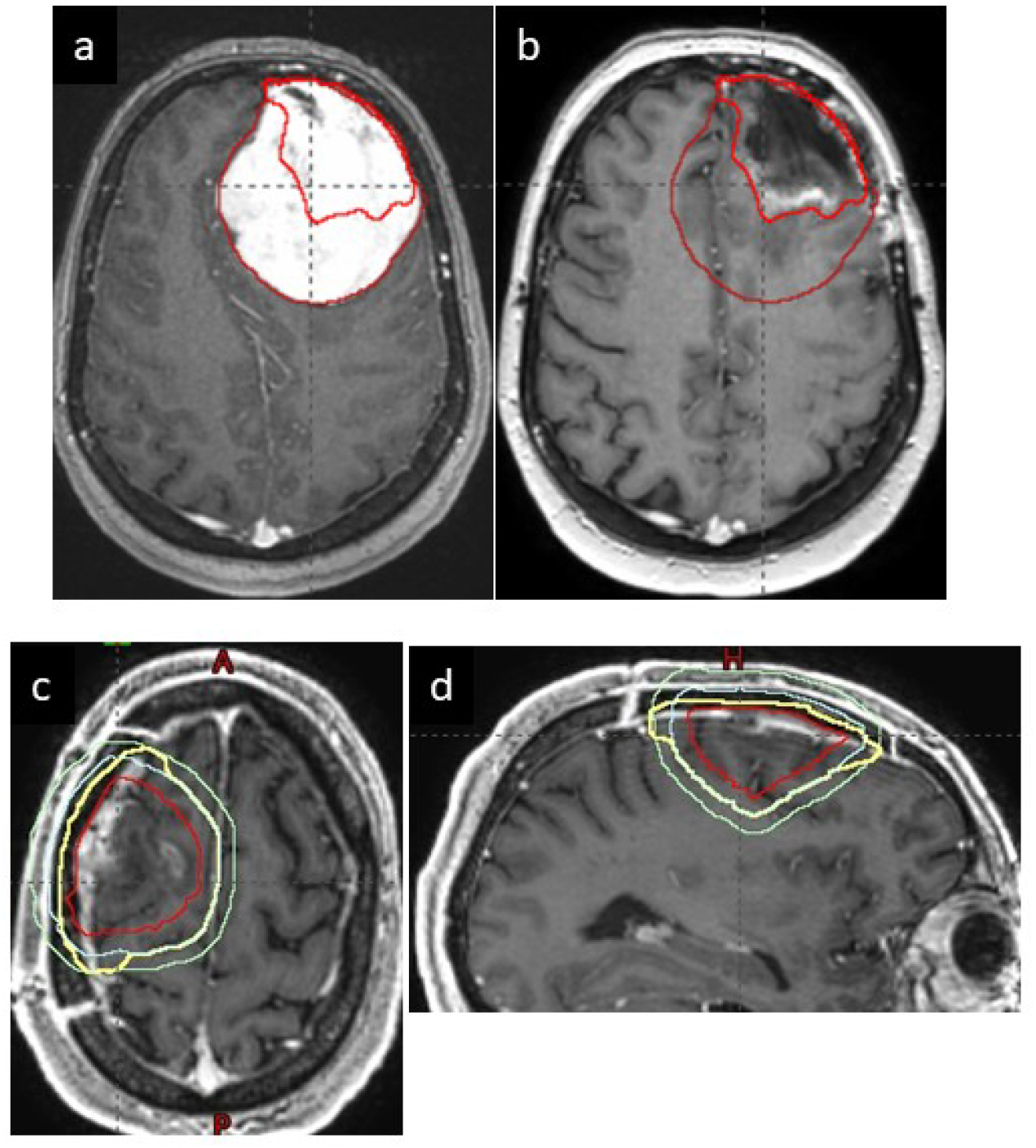
Patient A: Pre-op (a) and Post-op (b) MR’s showing how the GTV (bold / red) has been derived from the pre-op tumour (brown) but excludes the normal brain that has moved into the post-op cavity. Patient B: Transverse (c) and Sagittal (d) post-op MR showing how the CTV (bold / yellow) is derived from the GTV (red) by combining margins of 5mm (light blue) with 10mm (light green) along the dura.

For planning, they defined how to produce the planning target volume (PTV) (CTV with 5mm margin, reduced to 3mm for high precision techniques), how to deal with a superficial PTV by creating PTV_report, brought in by 5mm from the surface when optimising an inverse plan, which OAR were required and their dose constraints. They gave advice on how to treat conflicting situations where the PTV overlapped an organ at risk (OAR), stating the first priority was not to exceed the mandatory dose constraints for the optical tract and brainstem, the second was to ensure that the PTV (or PTV_report) was encompassed by the 95% isodose. Soon after the trial opened, UK RTTQA organised outlining and planning workshops for UK and European sites, where the radiotherapy protocol was explained. Centres were asked to bring outlined cases and were given feedback on their outlining technique.

All participating centres were credentialed. They had to complete an EORTC Facility Questionnaire and a Dummy Run (Completion of Benchmark cases for outlining and planning and Data transfer to QA team). The results were returned to the lead reviewing Centre for assessment and comments.

During the trial, ICR’s of outlining and planning were performed, prior to the start of radiotherapy treatment (i.e. prospectively) for all patients randomised to receive radiotherapy. Resubmissions and second reviews were required if a major variation was found (see assessment criteria below). Reasons for rejections were analysed and certain common themes emerged. This was reported to centres in Trial Newsletters and discussed at a number of centre visits organised for centres with less compliance and recruitment.

## Materials and Methods

In this paper, we use the written reports to analyse the outcome of the Quality Assurance process and to derive implications for future trials and wider radiotherapy delivery in the management of meningioma.

### Pre-Trial Radiotherapy Quality Assurance

Benchmark cases were assessed against pre-agreed standard outlines and plans. The pre-trial reviews were recorded on standardised reports which were returned to the centres. Detailed feedback was provided where necessary. Resubmissions were only asked for if there were gross variations from the guidelines leading the reviewers to be concerned that the centre hadn’t adequately understood the planning guidelines.

### On-Trial Radiotherapy Quality Assurance

In order to facilitate the ICRs, RTTQA physicists emailed centres as soon as they were notified that a patient had been randomised to radiotherapy, to explain the process to be followed. Centres submitted the anonymised patient structures, clinical history and MR and CT scans after the patient had been outlined and before planning. If major variations were found in the outlining, the centre was immediately contacted with an appropriate explanation of the changes required and asked to resubmit after editing. Otherwise, centres were contacted within 24 hours and told that they could proceed to planning. A written review report was emailed thereafter.

### Assessment Criteria

Variations from protocol were categorised as either major or minor. Major variations were those that, if used for treatment, would result in a possible effect on the treatment outcome or on side effects. Examples of major variations included an inadequate target volume resulting in a decreased Tumour Control Probability (TCP) or an excessive target volume likely to increase the Normal Tissue Complication Probability (NTCP). In the case of plans, an example would be: failing to adhere to a mandatory dose constraint without a reasonable explanation. Minor variations were commented upon for information and educational purposes. This included correct naming of outlines, delineation of the pre-operative GTV, 5 and 10mm construction outlines for margining to CTV (as in Figure 1), and editing of CTV off bone. For plans the list included lack of conformity of the 95% isodose to the PTV, missing or incorrect outlining of distant OAR. Borderline (bdl) variations were treated as minor for the pre-trial Benchmark cases, and major for the ICRs.

All variations, major or minor, were noted in standardised review reports.

### Post trial RT QA analysis

After recruitment had finished, the Benchmark and ICR reports were collated and analysed and the ICRs were independently audited. For this analysis where free comments had been used either instead of, or as well as completion of the tabular options, they were put into the categories shown in Tables 1-4. A random sample of 10% of the outlines and plans from UK, EORTC and TROG cases, were reviewed and assessed by an independent expert oncologist from the EORTC.

**Table 1.** Variations in Outlining of the Benchmark Case.

|  | UK | EORTC | TROG | Total | Resubmissions as<br>a % of Centres |
| --- | --- | --- | --- | --- | --- |
| No. of Centres | 20 | 25 | 11 | 56 |  |
| No. of Submissions | 23* | 27 | 12 | 62 |  |
| No. of Centres asked to Resubmit | 1 | 2 | 1 | 4 | 7% |
| No. of Centres with major variations | 4 | 3 | 1 | 8 | 13% |
|  |  |  |  | **Total** | **Total as % of**<br>**Submissions** |
| **MAJOR VARIATIONS** |  |  |  |  |  |
| GTV outlining | 1 | 3 | 1 | 5 | 8% |
| CTV – Dural coverage | 1+ 3bdl** | 1+ 5bdl** | 1+ 1bdl** | 3+9bdl | 5% + 15%bdl |
| **MINOR VARIATIONS** |  |  |  |  |  |
| Naming Convention | 4 | 12 | n/a | 16 | 30% |
| Pre-Op GTV not used | 10 | 13 | 1 | 24 | 39% |
| CTV not expanded with 5mm and<br>10mm constructions | 11 | 9 | 2 | 22 | 35% |
| Inadequate editing of CTV off<br>BONE | 6 | 1 | 2 | 9 | 15% |
| PTV (if present) manually edited<br>after expansion | 1 | 1 | 1 | 3 | 5% |
| Optical tract not continuous | 11 | 10 | n/a | 21 | 34% |
| Distant OAR's missing or incorrectly outlined | 7 | 9 | 7 | 23 | 37% |

*\*1 Centre had 2 changes of Principal Investigator*
*\*\*bdl = borderline*
*n/a: not recorded*

**Table 2.** Variations in Planning of the Benchmark Case.

|  | UK | EORTC | TROG | Total | Resubmissions as a % of Centres |
| --- | --- | --- | --- | --- | --- |
| No. of Centres | 20 | 25 | 11 | 56 |  |
| No. of Submissions | 21* | 32 | 17 | 70 |  |
| No. of Centres asked to Resubmit | 0 | 7 | 4** | 11 | 20% |
|  |  |  |  | **Total** | **Total as % of Submissions** |
| **MAJOR VARIATIONS** |  |  |  |  |  |
| CTV D99% < 95% | 11 | 22 | 4 | 37 | 53% |
| PTV D98% < 90% | 0 | 4 | 2 | 6 | 9% |
| Mandatory OAR dose constraints exceeded | 1 | 12 | 4 | 17 | 25% |
| CTV-PTV margins incorrect | 1 | 2 | 0 | 3 | 5% |
| PRV margins incorrect for OAR close to PTV | 1 | 3 | 0 | 4 | 6% |
| No. of plans that failed >1 mandatory dose constraint | 1 (5%) | 8 (25%) | 2 (12%) | 11 | 16% |
| **MINOR VARIATIONS** |  |  |  |  |  |
| PTV D98%<95% where avoidable | 4 | 12 | n/a | 16 | 23% |
| 95% isodose not well conformed to PTV | 10 | 13 | 1 | 24 | 35% |
| Optimal OAR dose constraints exceeded | 11 | 9 | 2 | 22 | 32% |
| Dose > 100% outside PTV | 6 | 1 | 2 | 9 | 13% |
| PTV_report incorrectly created or missing | 1 | 1 | 1 | 3 | 5% |
| Missing distant OAR | 11 | 10 | n/a | 21 | 30% |

*\* 1 centre submitted 2 plans for different planning systems*
*\*\*3 centres had 1 resubmission, 1 centre had 3 resubmissions*
*n/a: not recorded*

**Table 3.** Variations in Outlining of the Clinical cases.

|  | UK | EORTC | TROG | Total | Resubmissions as a % of cases |
| --- | --- | --- | --- | --- | --- |
| No. of Centres | 10 | 12 | 5 | 27 |  |
| No. of Clinical Cases | 30 | 27 | 7 | 64 |  |
| No. of Resubmissions | 13 | 16 | 2 | 31* | 48% |
|  |  |  |  | **Total** | **Total as % of clinical cases** |
| MAJOR VARIATIONS |  |  |  |  |  |
| GTV - inadequate outlining | 1 | 1 | 2 | 4 | 6% |
| CTV – inadequate coverage of Dura | 9 | 8 | 1 | 18 | 28% |
| CTV – Poor editing off Normal Brain | 2 | 4 | 0 | 6 | 9% |
| MINOR VARIATIONS |  |  |  |  |  |
| Naming Convention | 2 | 5 | 1 | 8 | 13% |
| Pre-Op GTV not used | 9 | 10 | 1 | 20 | 31% |
| CTV not expanded with 5mm and 10mm constructions | 2 | 1 | 2 | 5 | 8% |
| Inadequate editing of CTV off Bone | 1 | 2 | 1 | 4 | 6% |
| Optical tract not continuous | 4 | 3 | n/a | 7 | 11% |
| OAR's missing or incorrectly outlined | 8 | 10 | 2 | 20 | 31% |

*\*3 cases had 2 resubmissions*

**Table 4.**
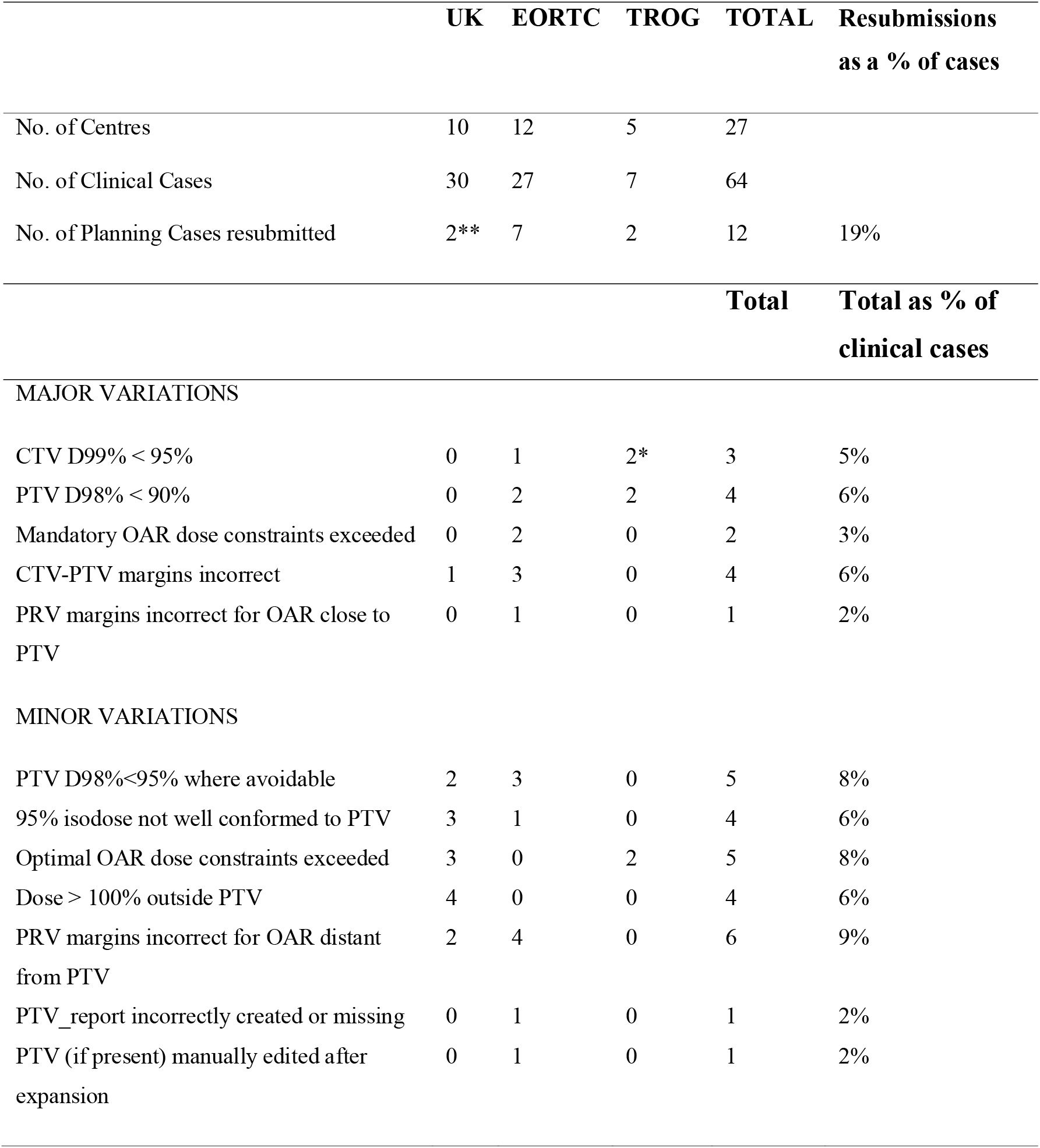
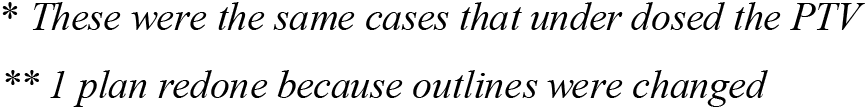
Variations in Treatment Plans of the Clinical cases.

|  | UK | EORTC | TROG | TOTAL | Resubmissions<br>as a % of cases |
| --- | --- | --- | --- | --- | --- |
| No. of Centres | 10 | 12 | 5 | 27 |  |
| No. of Clinical Cases | 30 | 27 | 7 | 64 |  |
| No. of Planning Cases resubmitted | 2** | 7 | 2 | 12 | 19% |
|  |  |  |  | **Total** | **Total as % of**<br>**clinical cases** |
| MAJOR VARIATIONS |  |  |  |  |  |
| CTV D99% < 95% | 0 | 1 | 2* | 3 | 5% |
| PTV D98% < 90% | 0 | 2 | 2 | 4 | 6% |
| Mandatory OAR dose constraints exceeded | 0 | 2 | 0 | 2 | 3% |
| CTV-PTV margins incorrect | 1 | 3 | 0 | 4 | 6% |
| PRV margins incorrect for OAR close to PTV | 0 | 1 | 0 | 1 | 2% |
| MINOR VARIATIONS |  |  |  |  |  |
| PTV D98%<95% where avoidable | 2 | 3 | 0 | 5 | 8% |
| 95% isodose not well conformed to PTV | 3 | 1 | 0 | 4 | 6% |
| Optimal OAR dose constraints exceeded | 3 | 0 | 2 | 5 | 8% |
| Dose > 100% outside PTV | 4 | 0 | 0 | 4 | 6% |
| PRV margins incorrect for OAR distant from PTV | 2 | 4 | 0 | 6 | 9% |
| PTV_report incorrectly created or missing | 0 | 1 | 0 | 1 | 2% |
| PTV (if present) manually edited after expansion | 0 | 1 | 0 | 1 | 2% |

*\* These were the same cases that under dosed the PTV*
*\*\* 1 plan redone because outlines were changed*

### Ethics Approval

The Newcastle & North Tyneside 2 Research Ethics Committee (15/NE/0013) gave ethical approval for this work. The trial was registered with <u>ISRCTN71502099</u>.

## Results

The trial opened on April 28^th^, 2016. One hundred and fifty-seven patients were recruited, and the last patient was recruited on May 21^st^, 2021. Fifty-six sites participated in the trial: 20 UK, 25 EORTC and 11 TROG.

All 56 Centres submitted the outlining and planning Benchmark cases. For the outlining cases, 52 (93%) were approved first time of which 12 (21%) centres had no variations and 31 (55%) only had minor variations. Eight Centres (13%) had 1 or 2 major variations. Of these, 4 (7%) centres were asked to re-submit and passed the resubmission.

The most common area of variation was the delineation of the GTV and CTV. Of note this included: failure to include the medial extent of involved dura in the GTV (best seen on the preoperative MRI); failure to include the most superior or inferior extent of involvement and inadequate dural margin for the CTV. Minor variations included lack of evidence that the CTV had been constructed according to the guidelines and failing to edit CTV from uninvolved bone. Many submissions had not outlined the optic nerves and chiasm as a continuous structure. This was classed as a minor variation as long as the Planning Risk Volume (PRV) (Optic tract with a 3mm margin) was continuous. The variations are summarised in Table 1.

For the planning cases, 56 centres submitted 57 plans (one for two planning systems), of which 45 (80%) were approved first time. For the benchmark plan the CTV was close to skin and to the optics PRV so the 2 most common major variations were insufficient coverage of the CTV with the 95% isodose and/or exceeding the mandatory dose constraint for the optical tract. Under coverage of the CTV or PTV near the surface did not automatically trigger a resubmission (as can be deduced from Table 2) unless combined with another major or several minor variations. The variations are summarised in Table 2.

Of the 56 activated centres, 27 centres treated patients with radiotherapy during the trial. Of the 78 patients randomised to radiotherapy, 64 (82%) patients received it: UK (30), EORTC (27) and TROG (7). Thirty-one (48%) sets of outlines (3 cases twice) and 12 (14%) treatment plans (1 plan twice) needed to be resubmitted (summarised in Tables 3 and 4).

For the outlining (Table 3), the most common major variation (28% of cases) was an inadequate coverage of the dura, especially at the dural attachments, when delineating the CTV and the most common minor variations were a failure to use the pre-op GTV to help outline the final GTV (31%), and inadequate outlining of OARs distant from the PTV.

For the plans, there were fewer major failures (Table 4). The two most common ones were inadequate coverage of the PTV with the 90% isodose (4 plans), and incorrect CTV-PTV margining (either using 3mm margins without adequate immobilisation or cropping the PTV manually (4 plans).

Patient outlines were reviewed and returned within one week of submission. Fifty-seven cases were submitted within the protocol timelines and radiotherapy was started on time. Three cases were submitted late with only one or two days for the review. Reviews were returned and approved for treatment within the protocol timelines. In four cases the outlines were only received after the patients had started radiotherapy. A retrospective review was performed.

The graph (Figure 2) of the number of patients per centre shows that 14 (50 %) of centres treated 1 patient within the trial. Eight (30%) of centres treated more than 2 patients within the trial. Two centres accrued 16 (25%) of all RT patients.

**Fig 2.**
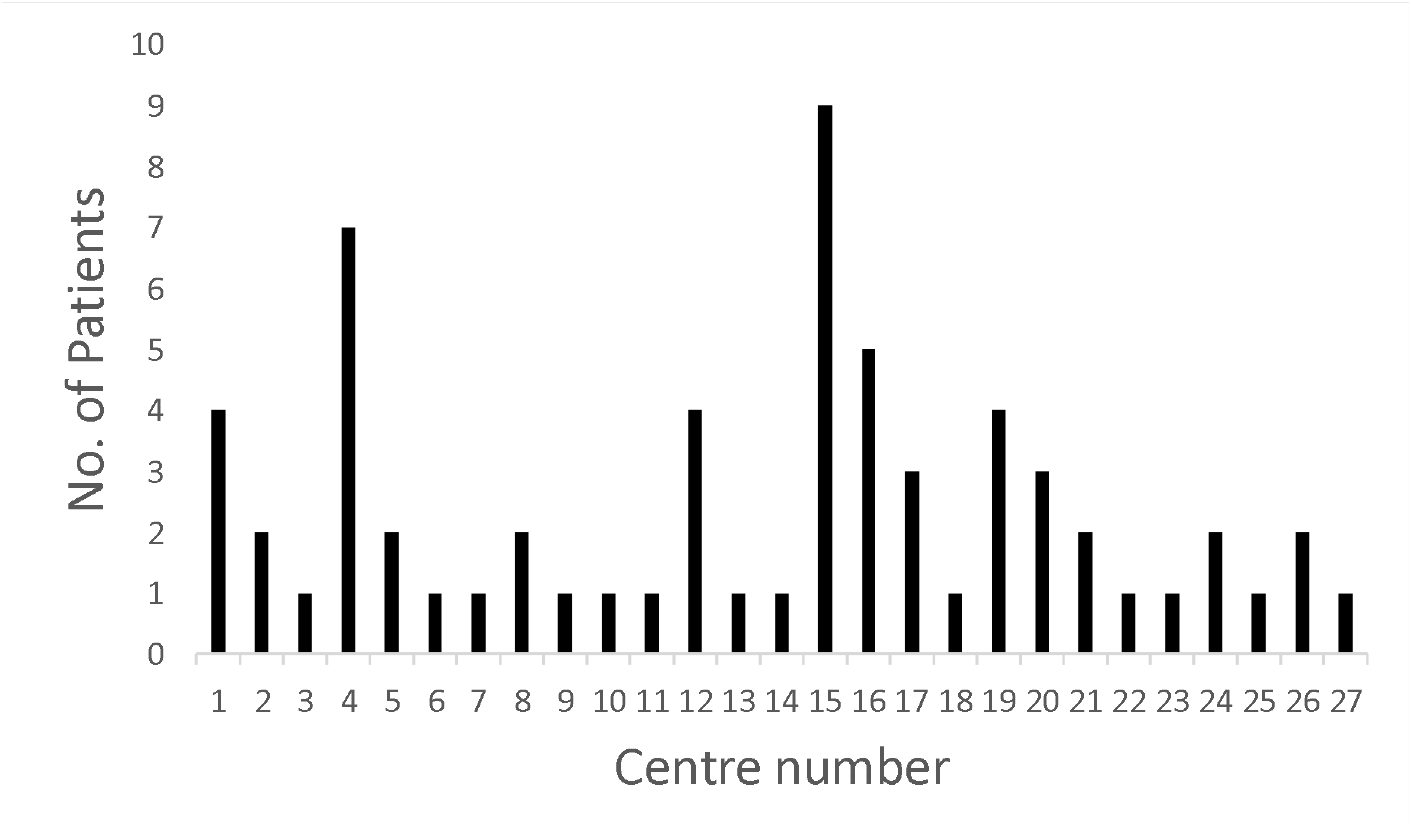
No. of Patients recruited per Centre.

## Discussion

The ROAM/EORTC-1308 trial undertook prospective RT QA to ensure consistent planning and delivery of radiotherapy to atypical meningioma. Similar to the RT QA review of the CATNON trial [11] we observed a substantial rate of non-compliance when reviewing the ‘on trial’ patients. Performing the reviews in advance of treatment enabled the non-compliances to be corrected before patients started radiotherapy.

Good communication is a key requirement for a successful trial. This is more challenging where international trials involve centres where there are several different “first languages”. The benchmark cases were a useful test of communication to ensure centres had understood the RT guidelines. The Dummy Run also tested whether data could be transferred efficiently between the Centre and the RTTQA group. For this trial the purpose of assessing the benchmark cases was primarily to check that the planning methods in the RT guidelines had been understood and that the communication system was rapid and robust. This is reflected in the small number of major variations that triggered a resubmission request. RT QA processes at the beginning of a trial, particularly requests for benchmark resubmissions, while important, can delay the opening of Centres, so instead, reports were provided with educational feedback. This decision was made in the knowledge that all trial patients would have their outlining and planning reviewed before treatment.

Doing prospective ICRs with limited core staff presented its own challenges. The system of emailing the staff in the Centres that were involved in planning to remind them that they would need to anonymise and send patient data as soon as it was ready and of phoning or emailing them immediately (rather than wait to produce the standard report) if there were concerns, was developed after reflecting on what improvements could be made after the first 10 patient reviews.

From feedback received, it was felt the workshops were a valuable tool in communicating the required Radiotherapy protocol. Physicists and radiographers were included in the site visits and were shown practical examples of radiotherapy outlining and planning.

The most common issue seen in target volume delineation was the failure to outline and submit the pre operative GTV (Table 3). It is possible that this had been referred to in the final post operative GTV outline, however where it was not submitted it was not possible to judge this. Often when a pre operative GTV was constructed at review it was clear that an involved dural margin had not been accounted for in the post operative outlining. Similarly, failure to include the 5 and 10mm construction lines made it difficult to understand how the final CTV had been constructed. In some cases, it was difficult to obtain a timely histological report to know whether there had been brain or bone involvement and hence what margin to use.

In general, the OARs were well outlined but in some cases the use of auto-outlining had not been corrected for, especially in contouring the Brainstem. One other common error was associated with outlining the optic apparatus and the position of the chiasm. It is possible this may in some cases be due to issues with registration and the transfer of images, however in others the optic apparatus was outlined as a discontinuous structure and raised the possibility of a hotspot arising in the sensitive optic nerve. In practice the effect of this was minimised in most cases by the construction of the PRV.

Atypical meningioma is a rare tumour. Only half of the participating centres had patients randomised to radiotherapy due to the low accrual rate for individual centres. The two centres that accrued approximately 25% of all RT patients had a resubmission rate of 25% (compared to 48% for the whole cohort) which demonstrates the benefit of learned experience. [12]

As with EORTC 22042 [14] there was no correlation between the number of variations in the Benchmark Cases and those in the ICRs. This is not surprising since we are dealing with a tumour where outlining and planning varies according to location in the Brain.

## Conclusions

To ensure consistent high-quality radiotherapy, prospective ICRs should be used for every patient in clinical trials of rare tumours where individual centres may only treat a few patients [13]. Pre-trial credentialing, with retrospective reviews of a sample of patients is not sufficient [14]. The RT QA process used in the ROAM/EORTC1308 trial should give assurance that the primary outcome (progression free survival: due to report in 2026) will be a robust assessment and any difference between treatment arms cannot be attributed to variation in radiotherapy planning or delivery.

## Data Availability

Individual participant data, after de-identification, will be made available beginning 9 months and ending 3 years after publication of the Article to researchers whose proposed use of the data is approved by the original study investigators. Proposals should be directed to the corresponding author and requesters will need to sign a data access agreement and comply with all applicable regulatory and governance requirements.

## Acknowledgements

The ROAM study is funded by the National Institute for Health Research Health Technology Assessment programme (NIHR-HTA) under grant agreement 12/173/14. The EORTC-1308 study is funded by contributions from the EORTC Brain Tumour group, the EORTC Radiation Oncology group and the University of Liverpool. UK RTTQA is also funded by NIHR.

The views expressed are those of the author(s) and not necessarily those of the NIHR or the Department of Health and Social Care

Supplementary Appendix A lists the contributors to the ROAM trial

## References

[1] Jenkinson MD, Weber DC, Haylock BJ, Mallucci CL, Zakaria R, Javadpour M. Atypical meningoma: current management dilemmas and prospective clinical trials. J Neurooncol. 2015;121:1–7.

[2] Aghi MK, Carter BS, Cosgrove GR, Ojemann RG, Amin-Hanjani S, Martuza RL, et al. Long-term recurrence rates of atypical meningiomas after gross total resection with or without postoperative adjuvant radiation. Neurosurgery. 2009;64:56–60; discussion

[3] Jenkinson MD, Javadpour M, Haylock BJ, Young B, Gillard H, Vinten J, et al. The ROAM/EORTC-1308 trial: Radiation versus Observation following surgical resection of Atypical Meningioma: study protocol for a randomised controlled trial. Trials. 2015;16:519.

[4] Keric N, Kalasauskas D, Freyschlag CF, Gempt J, Misch M, Poplawski A, et al. Impact of postoperative radiotherapy on recurrence of primary intracranial atypical meningiomas. J Neurooncol. 2020;146:347–55.

[5] Kessel KA, Fischer H, Oechnser M, Zimmer C, Meyer B, Combs SE. High-precision radiotherapy for meningiomas : Long-term results and patient-reported outcome (PRO). Strahlenther Onkol. 2017;193:921–30.

[6] Rebchuk AD, Alam A, Hounjet CD, Chaharyn BM, Gooderham PA, Yip S, et al. Survival and Recurrence Outcomes Following Adjuvant Radiotherapy for Grade 2 Intracranial Meningiomas: 13-Year Experience in a Tertiary-Care Center. World Neurosurg. 2022;161:e748–e56.

[7] Wujanto C, Chan TY, Soon YY, Vellayappan B. Should adjuvant radiotherapy be used in atypical meningioma (WHO grade 2) following gross total resection? A systematic review and Meta-analysis. Acta Oncol. 2022;61:1075–83.

[8] Zeng Q, Tian Z, Gao Q, Xu P, Shi F, Zhang J, et al. Effectiveness of Postoperative Radiotherapy in Patients with Atypical Meningiomas After Gross Total Resection: Analysis of 260 Cases. World Neurosurg. 2022;162:e580–e6.

[9] Goldbrunner R, Stavrinou P, Jenkinson MD, Sahm F, Mawrin C, Weber DC, et al. EANO guideline on the diagnosis and management of meningiomas. Neuro Oncol. 2021;23:1821–34.

[10] Simpson D. The recurrence of intracranial meningiomas after surgical treatment. J Neurol Neurosurg Psychiatry. 1957;20:22–39.

[11] Abrunhosa-Branquinho AN, Bar-Deroma R, Collette S, Clementel E, Liu Y, Hurkmans CW, et al. Radiotherapy quality assurance for the RTOG 0834/EORTC 26053-22054/NCIC CTG CEC.1/CATNON intergroup trial “concurrent and adjuvant temozolomide chemotherapy in newly diagnosed non-1p/19q deleted anaplastic glioma”: Individual case review analysis. RadiotherOncol. 2018;127:292–8.

[12] Trada Y, Kneebone A, Paneghel A, Pearse M, Sidhom M, Tang C, et al. Optimizing Radiation Therapy Quality Assurance in Clinical Trials: A TROG 08.03 RAVES Substudy. Int J Radiat Oncol Biol Phys. 2015;93:1045–51.

[13] Corrigan KL, Kry S, Howell RM, Kouzy R, Jaoude JA, Patel RR, et al. The radiotherapy quality assurance gap among phase III cancer clinical trials. Radiother Oncol. 2022;166:51–7.

[14] Coskun M, Straube W, Hurkmans CW, Melidis C, de Haan PF, Villa S, et al. Quality assurance of radiotherapy in the ongoing EORTC 22042-26042 trial for atypical and malignant meningioma: results from the dummy runs and prospective individual case Reviews. Radiat Oncol. 2013;8:23.

